# Pan-cancer Analyses reveal functional similarities of three lncRNAs across multiple tumors

**DOI:** 10.1101/2022.03.14.22272396

**Authors:** Abir Khazaal, Seid Miad Zandavi, Andrei Smolnikov, Shadma Fatima, Fatemeh Vafaee

## Abstract

Long non-coding RNAs (lncRNAs) are emerging as key regulators in many biological processes. The dysregulation of lncRNA expression has been associated with many diseases, including cancer. Mounting evidence suggests that lncRNAs are involved in cancer initiation, progression, and metastasis. Thus, understanding the functional implications of lncRNAs in tumorigenesis can aid in developing novel biomarkers and therapeutic targets. Rich cancer datasets, documenting genomic and transcriptomic alterations together with advancement in bioinformatics tools, have presented an opportunity to perform pan-cancer analyses across different cancer types. This study is aimed at conducting a pan-cancer analysis of lncRNAs by performing differential expression and functional analyses between tumor and normal adjacent samples across eight cancer types. Among dysregulated lncRNAs, seven were shared across all cancer types. We focused on three lncRNAs, found to be consistently dysregulated among tumors. It has been observed that these three lncRNAs of interest are interacting with a wide range of genes across different tissues, yet enriching substantially similar biological processes, found to be implicated in cancer progression and proliferation.

## Introduction

Cancer is a complex disease that continues to be a health burden globally [1]. It is characterised by dynamic alterations in the genome, including somatic mutations, epigenetic modifications, copy number variations and changes in expression profiles [2-4]. The emergence of massively parallel sequencing technologies has allowed for systematic documentation of the genetic changes in tumors and introduced the concept of the cancer genome [5-7]. Considered as a landmark cancer genomics program, The Cancer Genome Atlas (TCGA) program has produced, to date, more than 2.5 PB of multi layered genomic, transcriptomic, proteomic and epigenomic data along with clinical profiles for more than 11,000 patients, across 33 cancer types [8-10]. TCGA has improved our understanding of cancer genomics, revolutionised cancer classification and identified therapeutic targets [9,11,12].

Although cancers have their own genetic identity, with distinct, tissue specific changes, many tumors share similar genetic alterations that disrupt common biological processes [13,14]. Emerging computational technologies and rich datasets presented an opportunity to explore the differences and similarities of genetic and molecular changes across different tumor types using a set of techniques collectively referred to as pan-cancer analyses [14,15]. The importance of pan-cancer profiling lies in its ability to provide a comprehensive analysis of the genetic changes associated with multiple cancers. In addition, not only does it identify shared patterns, which aids in the development of uniform treatments strategies, but also distinguishes those unique alterations and enhances personalised care [14].

With the decreasing cost of whole-genome sequencing, there is a growing focus on performing pan-cancer analysis on non-coding regions of the genome. An increasing body of evidence suggests noncoding RNAs (ncRNAs) play an important role in biogenesis of cancer [16]. While some ncRNAs have been well studied, such as microRNAs [17], other types have been studied less extensively, including lncRNAs. LncRNAs are transcripts with a length greater than 200 nucleotides, exhibiting similar molecular characteristics as messenger RNAs (mRNAs) but lacking an appreciable potential to code for proteins [18,19]. Localised either in the nucleus or cytoplasm, lncRNAs form a complex network of interactions with DNA, RNA and proteins [20]. Although it is still debatable whether the majority of lncRNAs are simply transcriptional noise, some have been attributed with important, distinct biological roles. For example, lncRNA *XIST* is known to initiate silencing of the inactive X chromosome during X inactivation [21,22]. More generally, studies have suggested lncRNAs as cis- and trans-acting regulators of gene expression via chromatin reprogramming [23,24]. They have also been implicated in post-transcriptional regulation, including mRNA translation [25], as well as cell differentiation and development [26]. Despite these findings, lncRNA functions remain poorly understood. Nevertheless, lncRNAs are engaged in many processes and cellular functions; their dysregulation has been linked to many diseases, including cancer [16,27].

Aberrant expression of lncRNAs has been identified in many different tumors including brain, breast and colon cancer [28,29]. Many lncRNAs have also been shown to be regulated by oncogenes and tumor suppressors, suggesting a role in oncogenesis [30]. Furthermore, functional studies have revealed validated cancer roles for more than a hundred lncRNAs in tumors [31]. The discovery of lncRNAs has added another layer of complexity to cancer biology, with encouraging findings for potential clinical use.

A wide range of cancer treatments are currently available, including targeted drug therapy, radiotherapy, chemotherapy, laser therapy and surgery [32,33]. Chemotherapy, however, continues to be preferred despite the fact that its effectiveness diminishes when cancer has advanced or metastasized [33,34]. Poor prognosis is probably due to late diagnoses of cancer, together with tumors having acquired drug resistance, and continues to be a major challenge in treating malignancies [34]. It is thus important to search for new biomarkers for early diagnosis and therapeutic targets for more effective treatments. A plethora of evidence has revealed dysregulation of lncRNAs to be associated with cell proliferation, apoptosis and drug resistance, processes found implicated in the pathogenesis of cancer [35,36]. These findings put forward lncRNAs as potential biomarkers and therapy agents.

At present, there is an increasing focus on identifying lncRNAs associated with tumorigenesis and elucidating their functional implications. Rich RNA-seq datasets are a promising tool for this purpose, but their use can be computationally challenging. To highlight the importance of lncRNA association with cancer and overcome these challenges, The Atlas of Noncoding RNAs in Cancer (TANRIC) was developed [37]. TANRIC is a free and interactive database which gives users access to genomic, proteomic, clinical and lncRNAs expression data of 8,143 samples (tumorous and normal) from TCGA and others.

In order to characterise common, aberrantly expressed lncRNAs we performed a pan-cancer analysis on lncRNA expression profiles from TCGA derived dataset using TANRIC platform. We hypothesise that those found to be implicated in different cancer types may exhibit similar functional implications across cancers. To assess this hypothesis, we sought to identify dysregulated lncRNAs across eight TCGA cancer types and explored commonality among different malignancies. We then investigated their functional implications, by performing functional analysis and exploiting the similarity of enriched biological processes. Previous pan-cancer studies have focused on somatic mutations of whole genomes [15], tumor microenvironments [38], as well as proteomic profiles [39]. Putative functions have been studied for onco-lncRNAs dysregulated in multiple cancers [40] without focusing on common consistently dysregulated lncRNAs or exploring similarity of gene ontologies across different cancer types as performed in our study.

## Materials and Methods

An overview of the workflow of this study is shown in Figure 1 and explained in the following subsections.

**Figure 1.**
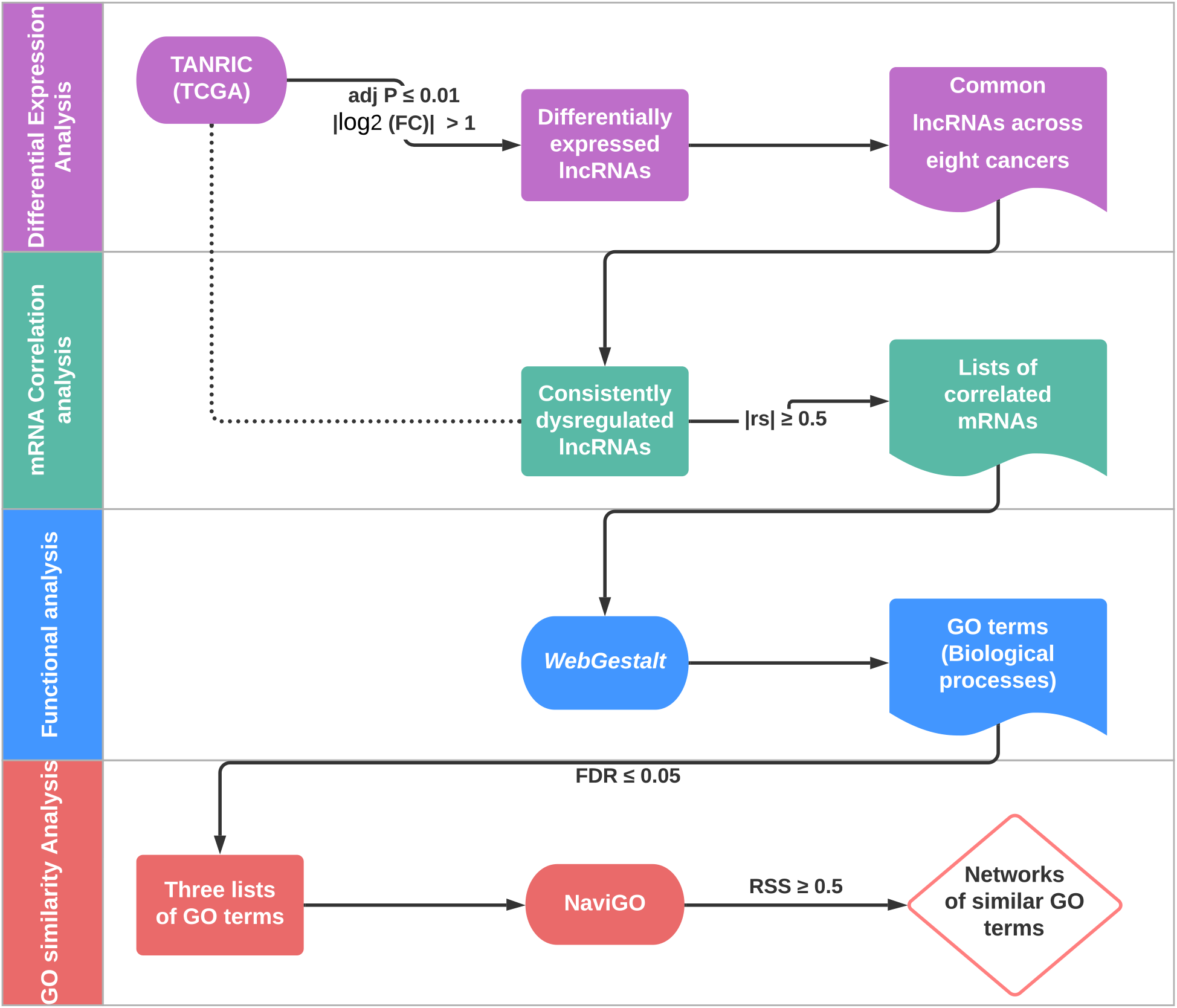
Overview of analyses performed in this study. The arrows show where data from an analysis has been used in the ensuing analysis

### Data source

We used TANRIC to access expression data of lncRNAs of 3,326 tumor samples and their related adjacent 416 normal tissue samples for eight cancer subtypes as categorised in TCGA. Included were 105 normal and 837 tumor samples of breast invasive carcinoma (BRCA); 58 normal and 488 tumor samples of lung adenocarcinoma (LUAD); 17 normal and 220 tumor samples of lung squamous cell carcinoma (LUSC); 52 normal and 374 tumor samples of prostate adenocarcinoma (PRAD); 59 normal and 497 tumor samples of thyroid carcinoma (THCA); 33 normal and 285 tumor samples of stomach adenocarcinoma (STAD); 50 normal and 200 tumor samples of liver hepatocellular carcinoma (LIHC) and 42 normal and 425 tumor samples of head and neck squamous cell carcinoma (HNSC). The choice of cancer types in this study was based on the availability of data on the corresponding adjacent normal samples.

### Differential exprssion analysis & commonality exploration

We carried out differential expression analysis by comparing lncRNA expression levels between tumor and related adjacent normal samples of a given set. Expression data for a total of 12,727 lncRNAs was downloaded from TANRIC v2.0, for each of the eight TCGA cancer types. Data files were stored as matrices with rows representing lncRNAs and columns representing samples. We then identified differentially expressed lncRNAs (up and down-regulated) with the threshold of |log_2_(FC)|>1. We used the Student’s t-test to calculate *p-*values and applied the Benjamini and Hochberg method to control the false discovery rate (FDR) [41]. Differentially expressed lncRNAs with adjusted *p*-value ≤ 0.01 were considered statistically significant. We then examined different pools of dysregulated lncRNAs across cancers to find commonalities. We identified common dysregulated lncRNAs (found in 2+ cancers) and unique dysregulated lncRNAs (specific to a given cancer type). Commonality was evaluated between each pair of cancers and represented by Jaccard index (*J*) [42]. These analyses were implemented in MATLAB with the code available at the GitHub repository (https://github.com/VafaeeLab/PanCancer-lncRNAs).

#### mRNA correlation analysis

We utilised TANRIC to explore mRNAs correlated with common dysregulated lncRNAs across each cancer type. Our approach was guided by the “guilt by association” principle which infers putative functions of a given gene (or, in this study, lncRNA) based on the functions of the genes it is co-expressed with [43]. Spearman rank correlation coefficient (rs) were calculated to examine correlation relationships between lncRNAs of interest and mRNAs expression. Lists of mRNAs correlating strongly with lncRNAs expression were extracted for each cancer type based on cut offs of rs ≥ 0.5 or rs ≤ - 0.5, for positive or negative correlation respectively, and correlation *p*-value ≤ 0.01.

#### Functional enrichment analysis

Exploration of large sets of genes can be achieved by organising them based on common functional features, such as biological pathways, molecular functions, or biological processes. One of the most widely used and standardised ways to understand genes and their products is to explore gene ontologies (GO) [44,45]. Thus, to investigate functional implications of lncRNAs of interest, we performed GO enrichment analysis with particular focus on biological processes. We exploited WebGestalt (WEB-based gene set analysis toolkit), to identify GO terms enriched by mRNAs lists, found to be correlated with common differentially expressed lncRNAs, with the aim of perusing their functional role as a set [46,47]. Statistical analysis of GO enrichment was performed using a Fisher’s exact test with Hypergeometric null distribution [48,49]. Significantly enriched GO terms were determined as FDR ≤ 0.05.

#### GO similarity analysis

GO, considered as a universal vocabulary, is structured as a hierarchical directed acyclic graph (DAG) where each node represents a class of gene function (GO term), and the connection between two GO terms indicates different relationships, such as “is a” or “part of”. This hierarchy allows exploring semantic similarities among enriched GO terms which could imply functional similarities between the associated genes [50,51]. Following the annotation of mRNAs lists by ontology, we merged GO terms enriched by each lncRNA across different cancers and distinguished two separate groups: GO terms enriched by positively correlated genes and GO terms enriched by negatively correlated genes. After additional filtering, based on FDR ≤0.05, GO terms were then further investigated. Over the past decade, several methods and platforms have been developed to examine semantic similarity between GO terms [52-54]. In the present study, we chose to measure closeness of GO terms using NaviGO, an interactive software which allows the retrieval of functional similarity scores and visualisation as networks [55]. From the six different scoring schemes offered by NaviGO, we relied on Relevance Semantic Similarity (RSS), which measures relative depth and rareness of the biological processes involved [50]. RSS ranges from 0 to 1, with 0 representing zero similarity and 1 indicating very high similarity. Functional similarity networks were created using the NaviGO visualiser based on threshold RSS ≥ 0.5.

## Results

### Common dysregulated lncRNAs

Differentially expressed lncRNAs (|log_2_(FC)| > 1 and FDR ≤ 0.01) were identified across each cancer (Table S1). In total, 9,616 lncRNAs manifested significant differential expression across cancers. Whilst similarity between pairs of cancers, represented by Jaccard index, appears low (Figure S1), collectively the number of shared lncRNAs of one cancer type with the remaining types is quite high, with overlap ranging from ∼80% to ∼97% (Table S2). Of the large number of lncRNAs found to be overlapping among different cancer pairs, seven were observed to be differentially expressed in all cancer types (Table S3). Following the examination of log_2_(FC) values; often the same lncRNA deemed upregulated in one cancer type can be found downregulated in another, or the other way around. Nonetheless, three lncRNAs were found to be consistently dysregulated across all cancers: ENSG00000235904 (*RBMS3-AS3*) (hereafter, “Antisense”) and ENSG00000261472 (Novel transcript) (hereafter, “Novel”) are both upregulated, and ENSG00000272455 (*MRPL20-DT*) (hereafter, “Divergent”) is downregulated (Table 1). For this reason, we chose to focus in the present study on the three consistently dysregulated lncRNAs: Antisense, Novel and Divergent. To infer putative functions, we investigated the correlation between these three lncRNAs and mRNA expression across each cancer. In total, 3,141 coding genes were selected (|rs| ≥ 0.5 and *p*-value ≤ 0.01), with 2,185 mRNAs found to be co-expressed with Antisense, 69 mRNAs for Novel and 1,026 mRNAs for Divergent (Figure 2). We found little overlap between correlated mRNAs across different cancers. It appears that for a given lncRNA, the group of co-expressed mRNAs is specific for each cancer type (Figure 3). Full list of correlated mRNAs with lncRNAs of interest across cancers can be found in Table S4.

**Table 1.** log_2_(Fold change) and FDR values of three lncRNAs, consistently dysregulated across all cancers.

| Cancer | ENSG00000235904 |  | ENSG00000261472 |  | ENSG00000272455 |  |
| --- | --- | --- | --- | --- | --- | --- |
|  | Antisense |  | Novel |  | Divergent |  |
|  | log<sub>2</sub> FC | FDR | log<sub>2</sub> FC | FDR | log<sub>2</sub> FC | FDR |
| BRCA | 3.45 | 4.61E-100 | 3.48 | 4.01E-105 | -1.91 | 4.39E-06 |
| HNSC | 1.69 | 3.47E-04 | 2.51 | 4.69E-17 | -3.21 | 5.20E-05 |
| LIHC | 1.64 | 6.88E-03 | 1.61 | 9.34E-03 | -4.15 | 6.81E-04 |
| LUAD | 1.91 | 3.57E-09 | 2.46 | 3.09E-22 | -3.11 | 4.32E-07 |
| LUSC | 2.41 | 6.87E-07 | 2.28 | 1.15E-05 | -4.11 | 6.89E-03 |
| PRAD | 2.41 | 6.22E-29 | 1.40 | 2.37E-03 | -2.16 | 3.85E-06 |
| STAD | 2.40 | 3.50E-09 | 1.64 | 7.28E-03 | -3.64 | 1.94E-03 |
| THCA | 1.40 | 5.77E-05 | 1.33 | 5.22E-04 | -1.69 | 7.50E-06 |

**Figure 2.**
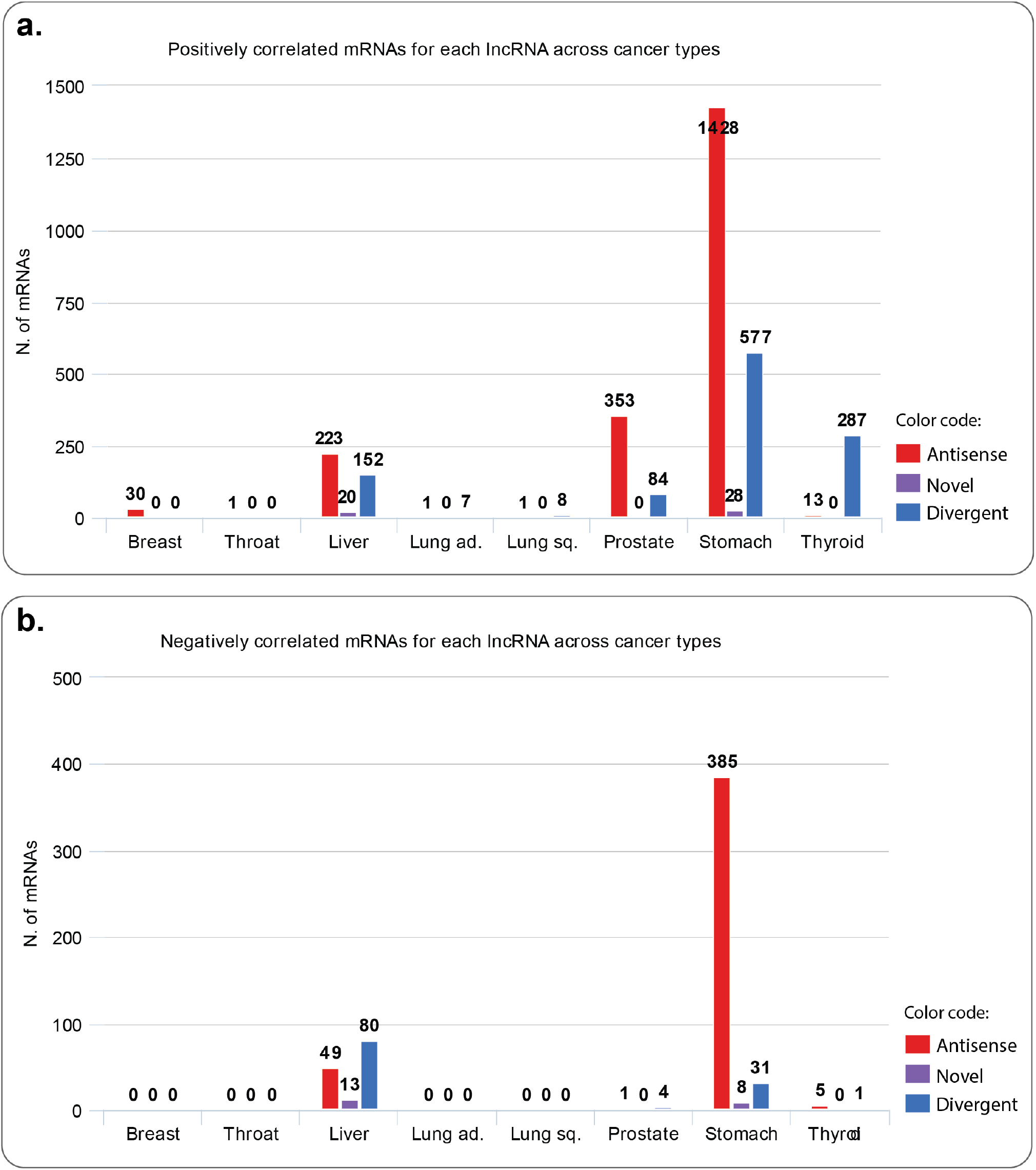
Bar plots showing the number of mRNAs related to the three lncRNAs, per cancer type. Red rep-resents Antisense, purple represents Novel and blue represents Divergent. **a**: number of positively correlated mRNAs. **b**: number of negatively correlated mRNAs.

**Figure 3.**
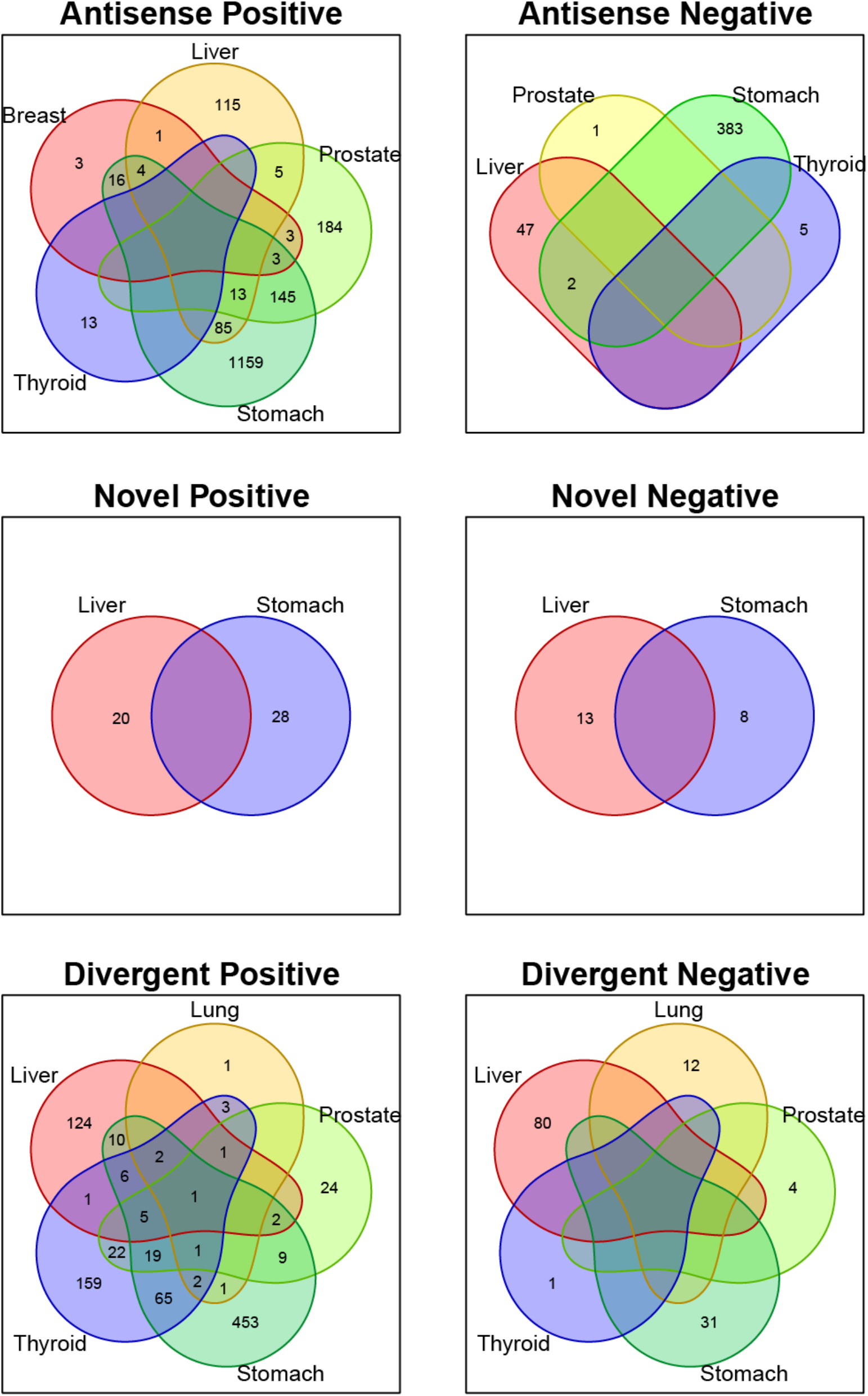
Venn diagram showing the overlap between lists of positively and negatively correlated mRNAs with lncRNAs: Antisense, Novel and Divergent, across different cancer types: Breast (BRCA), Thyroid (THCA), Stomach (STAD), Prostate (PRAD), Lung (LUAD, LUSC) and Liver (LIHC).

### Gene ontologies for inference of functional similarity

After identifying GO terms enriched by different sets of statistically associated mRNAs, we combined GO terms enriched by each lncRNA across all cancers, depending on the nature of correlation (positive and negative). In total, six groups of GO lists were surveyed: Antisense positive, Antisense negative, Novel positive, Novel negative, Divergent positive and Divergent negative. Additional filtering of enriched GO terms based on FDR ≤ 0.05, resulted in acquisition of three GO terms lists for further research. Two lists of GO terms associated with Antisense and one list with Divergent, with no records linked with lncRNA Novel (Figure S2). In order to provide functional elucidation of the remaining lncRNAs of interest, we explored similarity between GO term pairs using NaviGO and created GO pairwise similarity networks (RSS ≥ 0.05).

#### Similarity networks

Starting with GO list enriched by mRNAs positively correlated with Antisense, three clusters in the network of functionally similar GO terms were identified (Figure 4a). The first cluster contains GO terms predominantly involved in tissues and vessel morphogenesis, together with tissues and vasculature development (Figure 4b). The second cluster includes GO terms of system and cellular processes (such as actin-mediated cell contraction) in addition to localisation and movement of cell and/or subcellular component (Figure 4c). Finally, GO terms in the third cluster found of extracellular matrix and structure organisation along with biological and cell adhesion (Figure 4d). Network of mRNAs negatively correlated with Antisense, displayed GO terms appear to be mainly associated with ncRNA metabolic processes, particularly ribosomal RNA (rRNA) and ribosome biogenesis, in addition to ubiquitination (Figure 5). Lastly, mRNAs positively correlated with Divergent have enriched substantially similar GO terms, immersed with mRNA processing, splicing and metabolism, in addition to processes associated with cell cycle (Figure 6).

**Figure 4.**
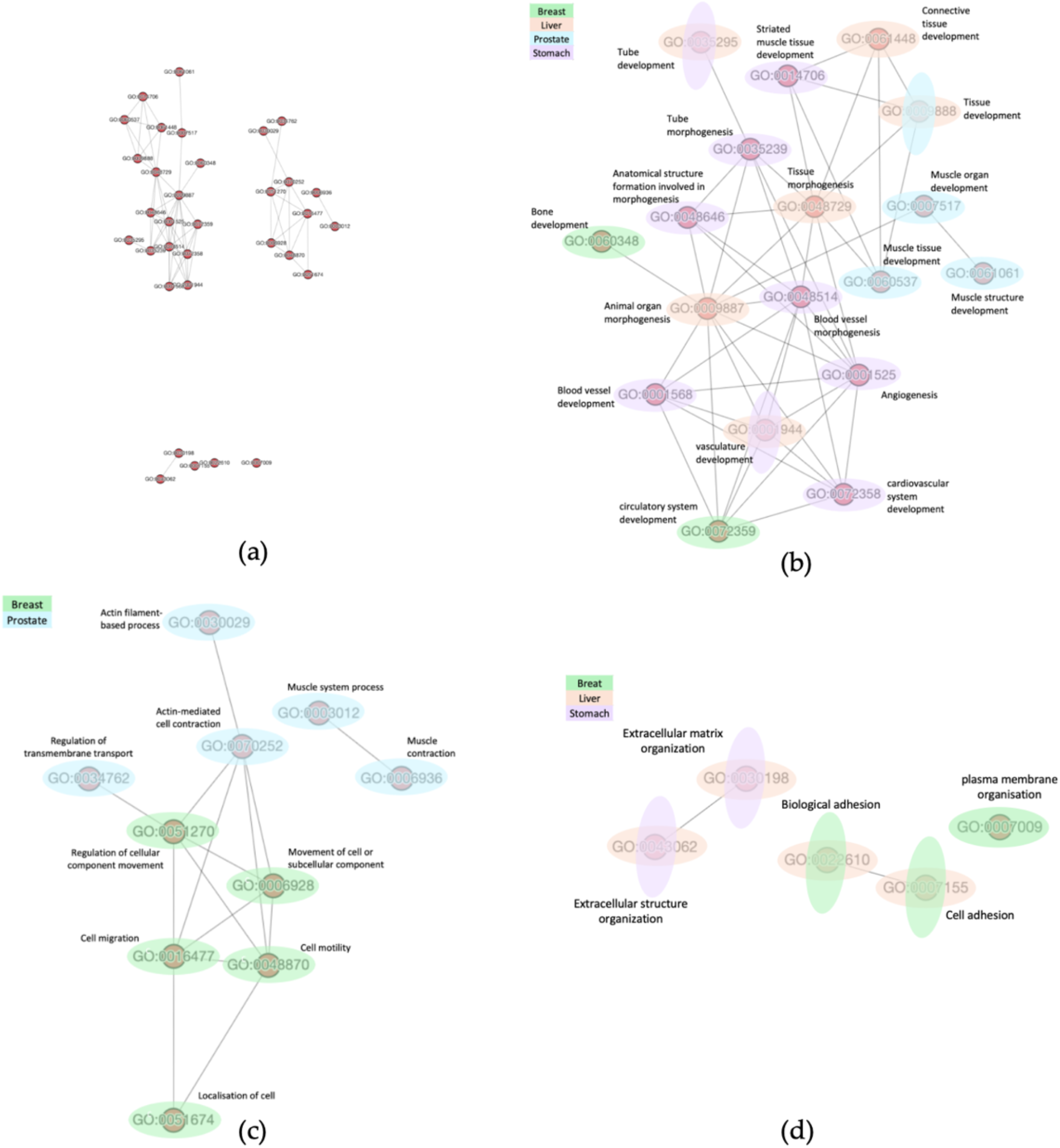
Functional similarity of GO terms associated with mRNAs of positive correlation with lncRNA Antisense. Network visualisation of functional similarity of GO terms enriched by positively correlated mRNAs with lncRNA “Antisense”, across cancers (when applicable, RSS ≥ 0.05). (**a**) Three clusters can be visually identified. (**b**) Close up of cluster 1. (**c**) Close up of cluster 2 (**d**) Close up of cluster 3. (**b**), (**c**) and (**d**): Each node indicates a GO term and the edges (lines in between) represent a functional similarity, representing relationships, such as “is a” or “part of”. The colour(s) of nodes represent the cancer type(s) in which GO term was found to be enriched, green for breast cancer (BRCA), orange for liver cancer (LIHC), blue for prostate cancer (PRAD) and purple for stomach cancer (STAD).

**Figure 5.**
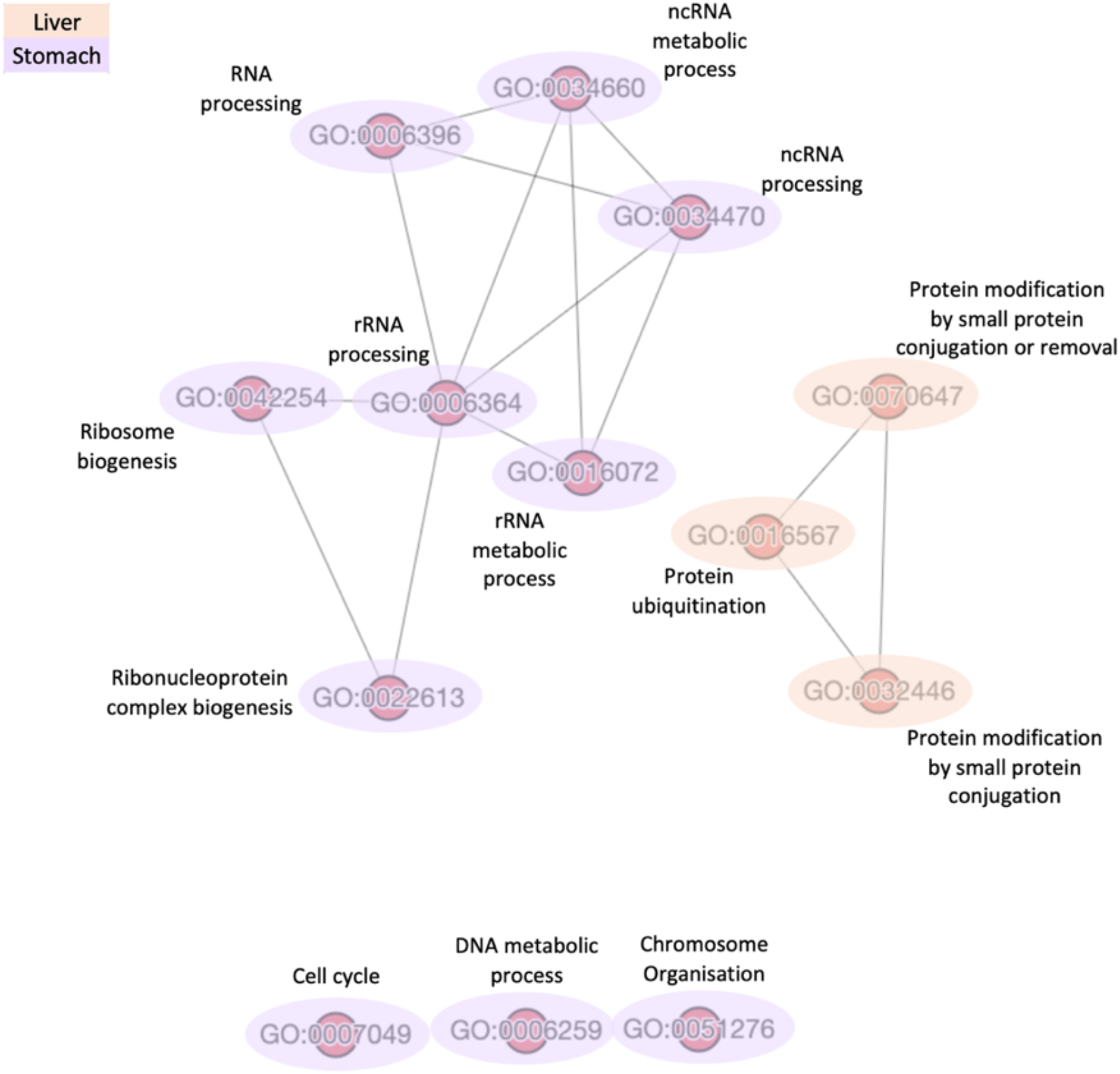
Functional similarity of GO terms associated with mRNAs of negative correlation with lncRNA Antisense. Network visualisation of functional similarity of GO terms enriched by negatively correlated mRNAs with lncRNA “Antisense”, across cancers (when applicable, RSS ≥ 0.05). The colour(s) of nodes represent the cancer type(s) in which GO term was found to be enriched, orange for liver cancer (LIHC) and purple for stomach cancer (STAD).

**Figure 6.**
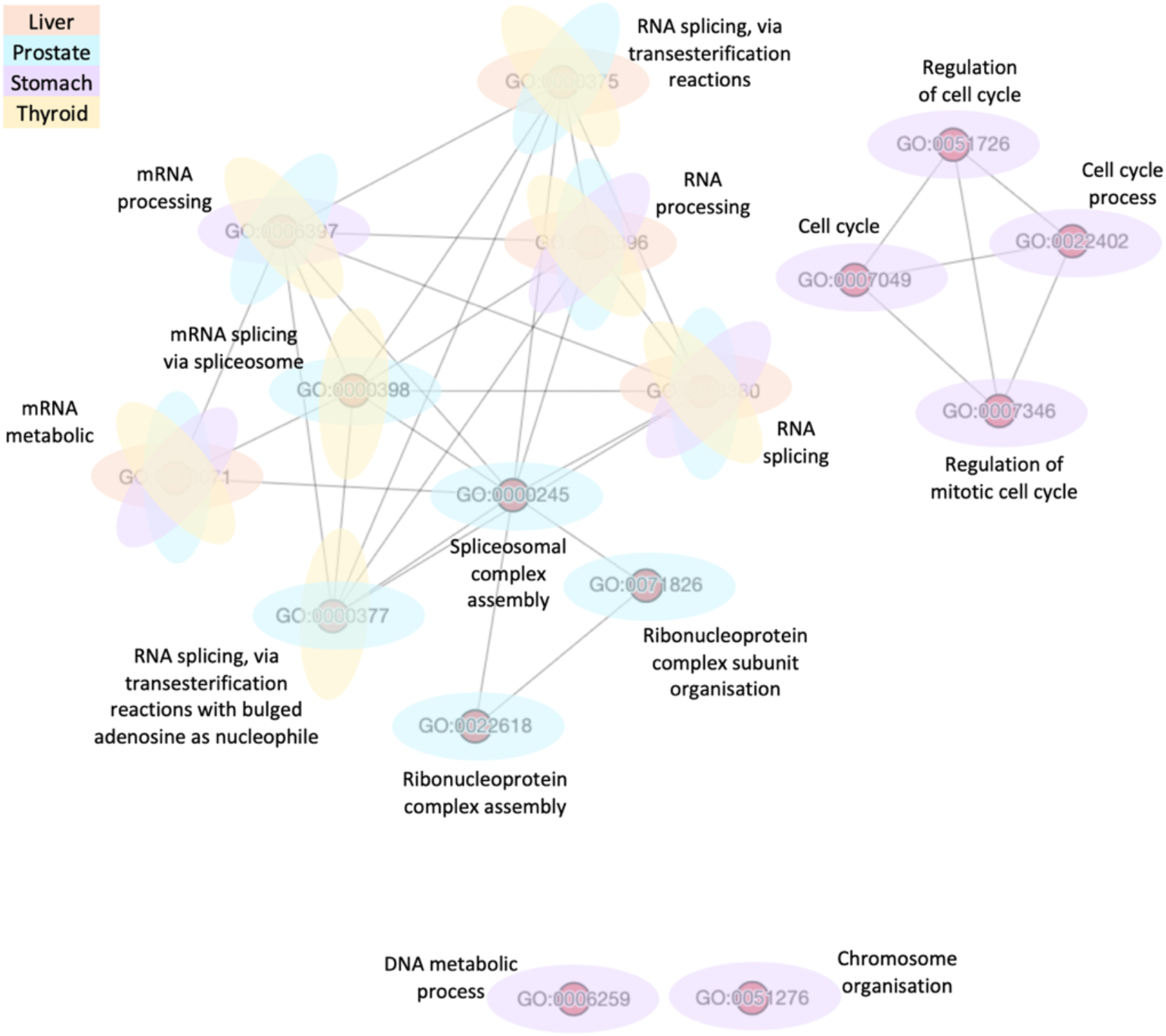
Functional similarity of GO terms associated with mRNAs of positive correlation with lncRNA Divergent. Network visualisation of functional similarity of GO terms enriched by negatively correlated mRNAs with lncRNA “Antisense”, across cancers (when applicable, RSS ≥ 0.05). The colour(s) of nodes represent the cancer type(s) in which GO term was found to be enriched, orange for liver cancer (LIHC), blue for prostate cancer (PRAD), purple for stomach cancer (STAD) and yellow for thyroid cancer (THCA).

Finally, we evaluated similarity scores of all GO terms enriched by both lncRNAs combined; comparison revealed high similarity between GO terms enriched by mRNAs negatively correlated with Antisense and those enriched by mRNAs positively correlated with Divergent (Figure 7). Interestingly, after further scrutinisation of the different networks, four GO terms were found to be shared between the two: DNA metabolic process, chromosome organisation, cell cycle and RNA processing (Figure 8).

**Figure 7.**
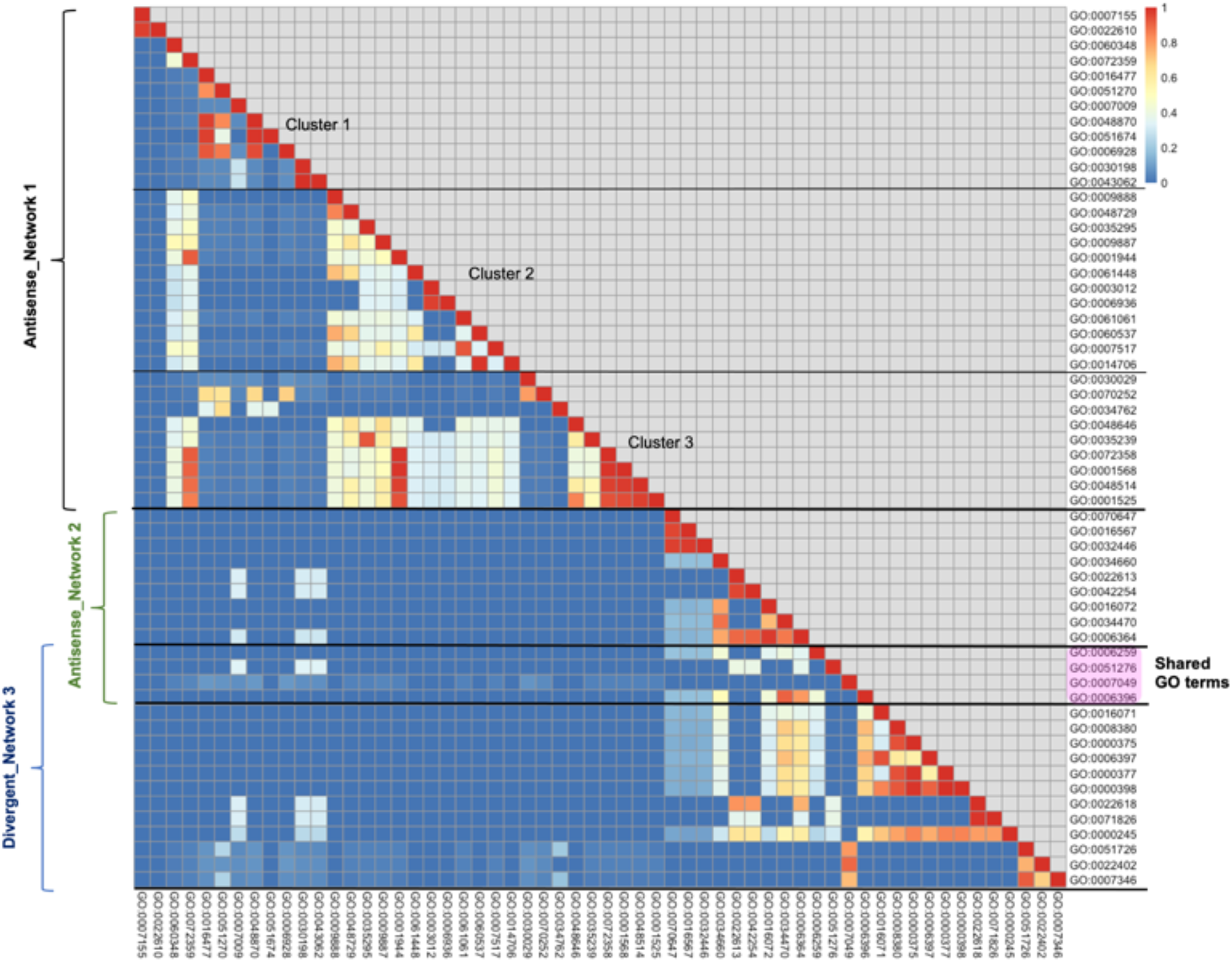
Summary of Semantic similarity scores of GO terms enriched by both lncRNAs “Antisense” and “Divergent”. Heatmap representing semantic similarity scores of GO terms enriched by all sets of mRNAs (combined), positively and negatively correlated (when applicable) with lncRNAs Antisense and Divergent. Blue indicates low similarity and red indicates high similarity. Three blocks can be seen. Three clusters can be identified in the first block, named Antisense Network 1. Shared GO terms between second block (Antisense Network 2) and third block (Divergent Network 3) are highlighted in pink.

**Figure 8.**
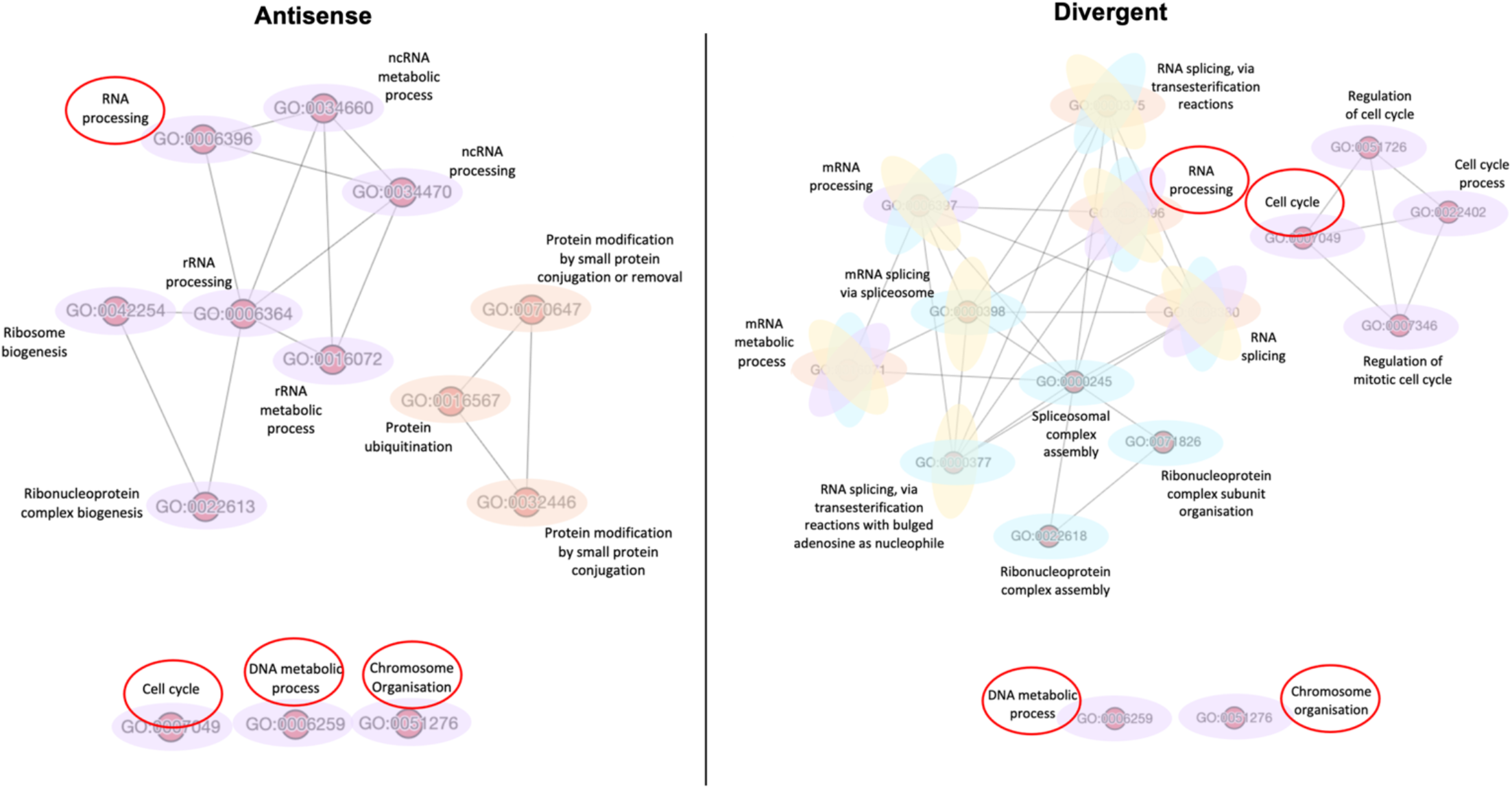
Comparison of Antisense and Divergent GO terms similarity networks. Similarity networks among GO terms enriched by mRNAs related negatively to Antisense (left) and mRNAs related positively to Divergent (right). Shared GO terms in both networks are encircled in red.

## Discussion

### Dysregulation of lncRNAs among cancers

The identification of 9,616 dysregulated lncRNAs suggests pervasive variation of lncRNA expression in cancers, consistent with previous studies [17,56]. Therefore, understanding functional implications of lncRNAs in malignancies is of high importance, as not only can it serve in developing diagnostic tools, it can also lead to new treatment strategies. Upon exploring commonality of dysregulated lncRNAs among different cancer types, it was observed that the number of common lncRNAs between tumors is quite high (average overlap ∼90%). These results suggest that potentially the same lncRNAs could be associated with different tumors across different tissues. Whilst it has been suggested that lncRNAs are cancer specific, displaying distinct expression patterns in different types of tumors, even at times within subtypes as well [56,57], some evidence show otherwise. For instance, *MALAT1* was suggested to be involved in multiple tumors; Inhibition of the well-studied lncRNA was found to prevent lung cancer metastasis [58]. Conversely, a more recent study showed that knocking out *MALAT1* actually promotes metastasis in breast cancer, suggesting its role as a metastasis suppressant [59]. Additionally, oncogenic role has also been proposed for *MALAT1* in colorectal carcinoma [60]. Nonetheless, we observed 2,855 lncRNAs to be dysregulated uniquely in one cancer type, denoting some level of specificity.

### Three consistently dysregulated lncRNAs

Intriguingly, seven lncRNAs were commonly dysregulated across all tumors, out of which three showed striking consistent dysregulation: ENSG00000235904, known as *RBMS3-AS3* gene and ENSG00000261472, a novel transcript, both exhibited up regulation in all tumor samples whilst ENSG00000272455, known as *MRPL20-DT* gene, manifested down regulation.

#### ENSG00000235904, RBMS3-AS3 “Antisense”

As with majority of lncRNAs, little is known about the functional implication of *RBMS3-AS3* or its association with tumors. According to lncATLAS [61], *RBMS3-AS3* is found to be expressed mainly in the cytoplasm. Generally, cytoplasmic lncRNAs are not well understood but believed, through formation of complexes with RNA binding proteins, to be involved in different mechanisms, such as mRNA translation and stability [62-64] and protein localisation [65,66]. With regards to cancer, *RBMS3-AS3* has been proposed as a competing endogenous RNA (ceRNA), targeted by several miRNAs in breast cancer [67]. In addition, *RBMS3-AS3* was shown to be serving as miRNA sponge, acting as a tumor suppressor in prostate cancer [68]. We explored TANRIC’s survival analysis for both of these cancer types, and found the survival rate across patients to be higher in those who have lower expression of *RBMS3-AS3* (Kaplan-Meier analysis and log-rank test, *p*-value < 0.05). Taken together, *RBMS3-AS3* seems to display aberrant expression patterns across tumors; further studies are required to investigate its function and potential involvement in cancer.

#### ENSG00000261472 “Novel”

Although this lncRNA was also found to be consistently upregulated across tumors, little is known about its association with cancer. ENSG00002614172 is small in size (<500bp), and unlike *RBMS3-AS3*, no localisation information was found in lncATLAS [61]. We explored its expression across tissues and found it to be almost negligible, with body fat having the highest value of only 1.7 TPM, according to the Genotype Tissue Expression project GTEX [69]. Lack of information on this transcript is possibly due to it being newly annotated, and most importantly minimally expressed across tissues. Searching through literature, we came across a breast cancer analysis where ENSG00000261472 was listed among other enriched lncRNAs [70]. However, to the best of our knowledge, no other studies have been published with reference to cancer. Concisely, ENSG00000261472 is a novel lncRNA whose cellular function is yet to be discovered.

#### ENSG00000272455, MRPL20-DT “Divergent”

Similar to Novel, no localisation information was detected in lncATLAS for *MRPL20-DT* [61]. Additionally, there seems to be no previous experimental studies investigating its function or possible involvement with tumor. However, a recent cancer analysis reported that the promoter of *MRPL20-DT* was among those who are consistently upregulated across 13 tumor types [71]. Likewise, a 2021 study communicated its upregulation amid other lncRNAs, in muscle invasive bladder cancer [72]. In contradiction, TANRIC’s survival analysis displayed better survival probability for those with higher expression of *MRPL20-DT* in bladder cancer (Kaplan-Meier analysis and log-rank test, *p*-value < 0.05), which comes in conformity with our results of it being downregulated in malignancy. In essence, *MRPL20-DT* role is still undetermined, but evidence suggest its possible association with cancer. Future research is needed to investigate its dysregulation in tumors and better understand the molecular mechanisms involved.

### Specificity of correlated mRNAs

In total, we found 3,141 mRNAs to be co-expressed with all three lncRNAs combined. Co-expressed gene lists, classified between positively and negatively correlated, ranged in size across different cancers and different lncRNAs, proposing a wide and diverse network of gene interactions across tumors. In addition, the number of correlated mRNAs of a given lncRNA was dependant on the tissue type. For instance, 1,428 mRNAs were positively correlated with Antisense in stomach cancer sample set, compared to only 353 and 30 mRNAs in prostate and breast cancer accordingly (Figure 2), suggesting some level of tissue specificity, which comes in accordance with previous findings [73]. Upon comparing different lists of mRNAs, although there is little intersect between malignancies, mRNAs were noted to be predominantly different (Figure 3), suggesting that, despite the commonality of these three transcripts, they appear to be interacting with different mRNAs in different tissues. Taken together, lncRNAs seem to manifest both tissue specific and ubiquitous relations, interacting with a broad range of genes across different tumors.

### Enriched GO and functional similarity

We investigated common functional features amongst statistically correlated gene lists by performing GO enrichment analysis, in order to understand the functional roles of lncRNAs of interest. We then differentiated three GO term lists, after further filtering by FDR. No GO terms were found to be notably associated with Novel post filtration, possibly because the number of correlated mRNAs was the lowest compared to Antisense and Divergent, hence, as a consequence no significant ontology enrichment was detected. The absence of GO terms is somewhat surprising, however does not undermine possible involvement of this lncRNA with tumors. It is worth noting that there is evidence of enrichment of Novel in breast cancer [70], in addition to the present study where consistent upregulation was outlined across all eight cancer types. These initial findings are promising; further studies would make a worthwhile contribution, to better understand the underlying mechanisms related to cancer.

### Networks of similar GO terms

Finally, to better understand the functional similarities of the two remaining lncRNAs Antisense and Divergent, we identified three GO similarity networks. It is worth noting that the scoring scheme (RSS) we adopted in creating these networks showed minimal variation when compared with Resnik and Lin’s semantic similarity, another two widely used measures [50,74]. Whilst this increases our confidence with results presented here, a caveat with this approach is that, network representation differs slightly based on the cut-off used with these scoring schemes. With that said, this is generally the case with many analyses and statistical tests which rely on arbitrary cut off values, and in this analysis, cut off value does not change the number or nature of biological processes involved, rather the way they are represented in a network.

#### GO terms of positively related mRNAs with “Antisense”

Three clusters of functionally similar GO terms can be identified in this network (Figure 4a). The first cluster comprised of biological processes linked to angiogenesis, blood vessel, tissue morphogenesis as well as vasculature development, all known to be critical for cancer growth (Figure 4b). For instance, it is well established that angiogenesis is one of the hallmarks of cancer [3]; Tumor cells recruit new blood vessels to allow for nutrients and oxygen delivery, as well to be able to metastasize to other tissues [75,76], and this also involves the development of new blood vessels and tissues. This is of importance as it implies that Antisense might be involved with pivotal mechanisms of tumorigenesis. Simultaneously, the second cluster encompassed GO terms of cell motility and migration, actin filament-based processes along with movement of cell (Figure 4c). These processes are also linked to those seen in the first cluster; taking cell motility for example, this is essential in allowing tumor cells to enter the vasculature, transport through blood vessels and invade other sites [77]. Moreover, networks of actin protein filaments form actin cytoskeleton, involved primarily in cell migration and motility in cancer, leading to metastasis [78,79]. Finally, the third cluster involved extracellular matrix and structure organisation together with biological and cell adhesion, processes also associated with cancer progression (Figure 4d). Indeed, there has been a focus on understanding the dysregulation of the extracellular matrix in complex diseases such as cancer. Being the major component of the tumor ‘microenvironment’, it has been suggested to modulate cell behaviour and influence adhesion and migration of cells [80,81]. Collectively, the GO terms presented in this network appear to be closely related, describing vital processes for the proliferation and progression of malignancies. The question remains though, whether Antisense is exerting a regulatory role in this network or is simply a by-pass product. Considering that accumulating evidence revealed many cellular functions to be regulated by lncRNAs [82], further investigation is required to understand the potential role of Antisense. As not only it will aid in understanding cancer pathogenesis, but could also be a promising target to enhance the efficacy of current therapeutic approaches, such as anti-angiogenic drugs [83].

#### GO terms of negatively related mRNAs with “Antisense”

The smaller list of negatively correlated mRNAs with Antisense enriched important functions relating to ncRNA metabolic processes, particularly rRNA, ribosome biogenesis and ubiquitination (Figure 5). Increased ribosomal biogenesis has been associated with tumor proliferation, but mounting evidence suggests that impaired ribosomal activity also drives tumorigenesis [84,85]. Ribosome biogenesis is an important regulator of cellular activities, including cell growth and cell cycle progression [86,87]; an increase in rRNA processing is observed during G1 of interphase, in preparation for protein translation, whilst during mitosis, downregulation of ribosomal activity is needed to signal the ending of cell cycle. Uncontrolled cell proliferation, a common feature in cancer, is a consequence of impaired ribosomal activity. Furthermore, it is now believed that perturbation of ribosomal biogenesis is sufficient to lead to malignant transformation [88]. In summary, imbalance and perturbation of ribosomal biogenesis is predicted to be linked with malignancies.

Another process found in this network is ubiquitination (also known as ubiquitylation), a post-translational mechanism in which proteins are tagged by the conjugation of ubiquitin, for modification. Ubiquitin is a small regulatory protein that is highly conserved in eukaryotes, most commonly found to initiate proteins degradation, apart from also altering protein-protein interactions and modulating cellular processes such as cell cycle, apoptosis, cell signaling and DNA repair [89,90]. It has been shown that cytoplasmic lncRNAs interfere with protein expression by either obstructing or promoting ubiquitination [91]. With a balanced ribosomal genesis for instance, tumor suppressor protein p53 is usually post-translationally downregulated through ubiquitination. However, studies have revealed that disruption of ribosomal biogenesis primarily causes activation of p53, and consequently disrupts its degradation through ubiquitination [84]. Hence, it comes as no surprise that these processes are interconnected, and their perturbation is associated with carcinogenesis. lncRNA Antisense, identified in this study, has been shown to be associated with a range of important biochemical processes, explicitly ubiquitination. Currently novel strategies are being developed to target certain pathways in which ubiquitin is primarily involved, resulting in potentiation of drug efficacy and overcoming multi drug resistance [92].

#### GO terms of positively related mRNAs with “Divergent”

Finally, the network of similar GO terms of mRNAs positively related with Divergent displayed processes, enriched in more than one cancer type, relating to mRNA processing, in particular splicing pathways in addition to cell cycle regulation, specifically mitosis (Figure 6). Accounting primarily for protein diversity, splicing is a fundamental step of mRNA processing where the same coding gene can have different, even at times, opposing functional transcripts called isoforms. It has been revealed that aberrant splicing is linked with cancer, yielding cancer specific isoforms favourable for tumor growth [93,94]. In addition, defective splicing also perturbs the cell cycle, which is also enriched in this network. Previous studies have shown a strong connection between several lncRNAs and cell division. For instance, oncogenic role has been suggested to lncRNA *EPIC1* as induced cell cycle arrest was detected upon its depletion [95]. Taken together, Divergent seem to be directly associated with pivotal processes, involved in cancer growth and development. Consistent with literature, lncRNAs have been suggested to influence splicing in diseases such as cancer [96]. Moreover, new mechanism of cell death modulation has also been linked to lncRNA through interaction with protein factors, leading to apoptosis resistance [97]. However, the downfall of these findings, is that the underlying mechanisms still remain largely unknown. Nonetheless, Divergent has been presented in this study to be consistently downregulated across tumors; investigating its possible role in splicing events and cell growth, may be of benefit.

#### GO terms shared across networks

We reported four processes to be shared by both lncRNAs (Figure 8). Interestingly, these processes were found in the network of negatively correlated mRNAs with Antisense, reported to be upregulated in cancers, and that of positively correlated mRNAs with Divergent, found to be downregulated in cancers. Taken together, it appears that although these two lncRNAs are interacting with different sets of mRNAs across different cancers, and possibly through different mechanisms with one being upregulated and the other shown to be downregulated; they are both enriching substantially similar processes, found to be fundamental in cancer proliferation and progression. Further research is required to identify whether consistently dysregulated lncRNAs are involved as causal agents of these processes, or simply a consequence of malignant tumor formation.

### Future direction

The foundational processes identified in this study, such as angiogenesis, underly all tumors regardless of tissue type. Understanding the role of lncRNAs in enriching these perturbations is thus very important and can be advantageous, particularly when malignant cells spread to other tissues, leading to current treatment strategies in becoming somewhat ineffective. A possible future direction would be to use other computational approaches and databases to decipher putative functions of lncRNAs, as well as using other data sources. For instance, taking an integrative approach considering transcriptomic and epigenomic data (e.g., methylation profile [98]), or genomic changes such as copy number alterations (CNAs) [99]. In addition, using UCSC genome browser [100], we could also examine the genomic location of lncRNAs along with co-expressed coding genes in order to explore cis and trans-relationships, which can be an initial step in discovering potential regulatory roles. Moreover, it would be feasible to investigate if those identified in this study are also dysregulated in other types of cancer. Furthermore, we have focused in the present study on commonly dysregulated lncRNAs across cancers. The complimentary, future approach could be to explore lncRNAs that are specific to each cancer type/subtype, with the aim of investigating the cancer specific functions that have been carried out by different lncRNAs, which can also be of benefit.

## Conclusion

Once seen as transcriptional by-products, lncRNAs are now emerging as key players in cellular function, regulating a wide range of biological processes, and involved in their disruption. LncRNA analysis is a rapidly developing field in cancer pathogenesis, with many lncRNAs found to be deregulated in tumors. Cancer is complex in nature; Despite tremendous efforts and progress in treatment strategies, poor prognosis continues to be an issue particularly when cancer has advanced or metastasized. Therefore, identifying novel biomarkers and treatment strategies is of focus, to improve early diagnosis and the prognosis of cancer patients.

Our study provides evidence that lncRNAs may be contributing to hallmarks of cancer, regardless of cancer type. It was observed that consistently deregulated lncRNAs across cancers are interacting with a wide range of protein coding genes, mostly shown to be specific to tissue type, but enriching substantially similar biological processes implicated in carcinogenesis. These results are promising, suggesting that lncRNAs can serve as potential therapeutic targets to be applied in multiple cancer subtypes. Future treatment strategies may potentially include non-coding genes in addition to specific protein targets.

Although a lot of progress has been made, only a handful of lncRNAs have had functional characterisation roles. The rapidly growing catalogue of cancer associated lncRNAs, led by advance of sequencing methodologies and computational tools, has posed a challenge to experimental validation. Thereby, new *in silico* approaches have emerged to aid in inferring functional roles for uncharacterised lncRNAs. LncRNAs have brought a promising new era to cancer biology, especially in terms of diagnosis and therapy.

## Supporting information

Lists of dysregulated lncRNAs

Lists of correlated mRNAs

## Data Availability

All data produced in the present work are contained in the manuscript

https://ibl.mdanderson.org/tanric/_design/basic/main.html

## Data Availability Statement

Data used in this study are publicly accessible via TANRIC, The Atlas of non-coding RNA in Cancer (www.tanric.org). Codes generated in this study are available at the GitHub repository https://github.com/VafaeeLab/PanCancer-lncRNAs.

## Author Contributions

Conceptualization, F.V.; methodology, F.V.; software, F.V.; formal analysis, S.M.Z.; investigation, A.K.; resources, F.V., A.K. and S.F.; writing—original draft preparation, A.K.; writing— review and editing, F.V. and A.K.; visualization, F.V. and A.K; supervision, F.V.; project administration, A.K.; funding acquisition, F.V. All authors have read and agreed to the published version of the manuscript.

## Acknowledgments

We wish to record our deep sense of gratitude and profound thanks to our research supervisor Dr. Fatemeh Vafaee for her guidance and constant encouragement. We also want to thank our colleagues in Vafaee laboratory at UNSW, BABS for their support and insightful comments with this project. Finally, AK wants to acknowledge the contribution of her partner, Rabeeh Krayem, who has supported her intellectually and emotionally throughout this study.

## Conflicts of Interest

Authors declare no conflicts of interest.

## Funding

This research received no external funding.

## Notes

### Competing Interest Statement

The authors have declared no competing interest.

### Funding Statement

This study did not receive any funding

