## Supplemental Information for "Pan-cancer Analyses reveal functional similarities of three lncRNAs across multiple tumors"

### List of Supplementary Figures and Tables:

**Figure S1.** Heatmap representing commonality of lncRNAs between different pairs of cancers.

**Figure S2.** Summary of gene ontologies (FDR ≤ 0.01) enriched across all cancers by each lncRNA.

**Table S1.** Lists of differentially expressed lncRNAs (|log2(FC)| > 1 and FDR ≤ 0.01, comparing tumors with adjacent normal samples) identified across each cancer.

**Table S2.** Distribution of differentially expressed lncRNAs (upregulated and downregulated) across TCGA cancer types.

**Table S3.** Details on lncRNAs found to be dysregulated across all cancer types.

**Table S4.** Lists of correlated mRNAs with three consistently dysregulated lncRNAs, across each cancer type**.**

| Cancer types | BRCA | HNSC | LIHC | LUAD | LUSC | PRAD | STAD | THCA |
|---|---|---|---|---|---|---|---|---|
| BRCA | | 0.19 | 0.23 | 0.31 | 0.14 | 0.23 | 0.17 | 0.31 |
| HNSC | 0.19 | | 0.17 | 0.23 | 0.13 | 0.18 | 0.20 | 0.16 |
| LIHC | 0.23 | 0.17 | | 0.26 | 0.08 | 0.22 | 0.23 | 0.23 |
| LUAD | 0.31 | 0.23 | 0.26 | | 0.23 | 0.23 | 0.22 | 0.25 |
| LUSC | 0.14 | 0.13 | 0.08 | 0.23 | | 0.12 | 0.09 | 0.12 |
| PRAD | 0.23 | 0.18 | 0.22 | 0.23 | 0.12 | | 0.18 | 0.22 |
| STAD | 0.17 | 0.20 | 0.23 | 0.22 | 0.09 | 0.18 | | 0.16 |
| THCA | 0.31 | 0.16 | 0.23 | 0.25 | 0.12 | 0.22 | 0.16 | |

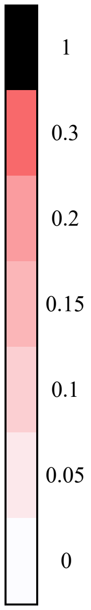


**Figure S1.** Heatmap representing commonality of lncRNAs between different pairs of cancers. Similarity scores amid pairs of cancers indicated by Jaccard index. *J* scores range from 0-1 with white shaded boxes (J=0) indicating low similarity and black shaded boxes (J=1) indicating very high similarity (identical sets of lncRNAs).

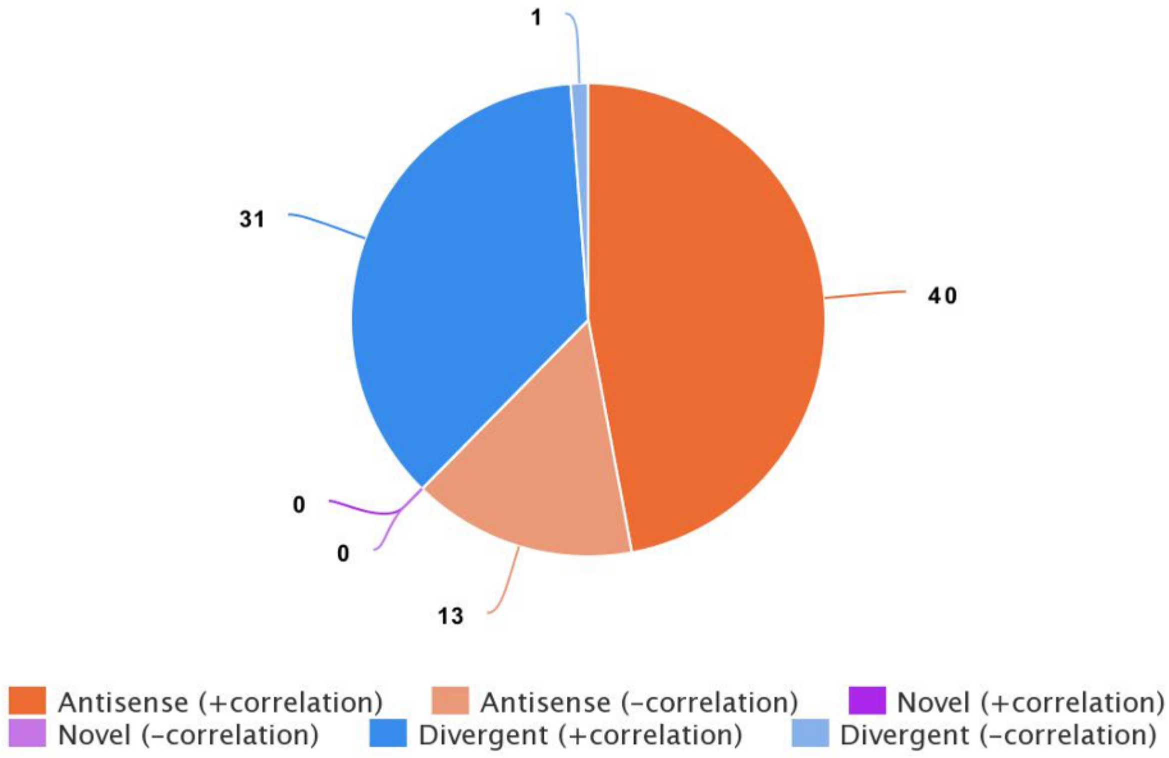


**Figure S2.** Summary of gene ontologies (FDR ≤ 0.05) enriched across all cancer types by each lncRNA: Antisense, Novel and Divergent. GO terms grouped by the nature of relationship with respective lists of mRNAs; positive and negative correlation.

**Table S1.** Lists of differentially expressed lncRNAs (|log2(FC)| > 1 and FDR ≤ 0.01), comparing tumors with adjacent normal samples) identified across each cancer.

*[Attached as an Excel Sheet]*

**Table S2.** Distribution of differentially expressed lncRNAs (upregulated and downregulated) across TCGA cancer types.

| Cancer | Breast (BRCA) | Throat (HNSC) | Liver (LIHC) | Lung (LUAD) | Lung (LUSC) | Prostate (PRAD) | Stomach (STAD) | Thyroid (THCA) |
|---|---|---|---|---|---|---|---|---|
| N. of differentially expressed lncRNA | 4807 | 2098 | 3024 | 3747 | 1212 | 2653 | 2101 | 4861 |
| % of upregulated lncRNAs | 69.8% | 50% | 5.8% | 47.8% | 93.8% | 57.3% | 21.3% | 81% |
| % of downregulated lncRNAs | 30.2% | 50% | 94.2% | 52.2% | 6.2% | 42.7% | 78.7% | 19% |
| % of common lncRNAs | 84.6% | 90.9% | 92.4% | 92% | 97.3% | 92% | 91.2% | 80.3% |

**Table S3.** Tables showing details on lncRNAs found to be dysregulated across all cancer types.

| Ensembl ID | Gene Name | Annotation | Location (GRCh38) | Strand | Transcripts |
|---|---|---|---|---|---|
| ENSG00000223561.7 | Novel | TAGENE | Chr7: 25,593,304-25,751,032 | reverse | 25 |
| ENSG00000257167.2 | TMPO-AS1 | Manual (Havana project) | Chr12: 98,512,973-98,516,422 | reverse | 2 |
| ENSG00000249859.12 | PVT1 | Manual (Havana project) | Chr8: 127,794,526-128,187,101 | forward | 176 |
| ENSG00000245522.2 | LINC02709 | Manual (Havana project) | Chr11: 9,754,770-9,759,533 | reverse | 2 |
| ENSG00000235904.3 | RBMS3-AS3 | Manual (Havana project) | Chr3: 29,054,570-29,290,726 | reverse | 5 |
| ENSG00000261472.1 | Novel | Manual (Havana project) | Chr16: 79,505,603-79,516,293 | forward | 1 |
| ENSG00000272455.1 | MRPL20-DT | Manual (Havana project) | Chr1: 1,409,096-1,410,618 | forward | 1 |

**Table S4.** Lists of correlated mRNAs (|rs| ≥ 0.5 and *p*-value ≤ 0.01) with three consistently dysregulated lncRNAs, across each cancer type**.**

*[Attached as an Excel Sheet]*
